# Multiple Psychiatric Traits Enriched for Brain Tissues in the Early Postpartum Period

**DOI:** 10.1101/2025.09.07.25335185

**Authors:** Natasha Berthold, Shuyang Yao, Jerry Guintivano

## Abstract

**Background:** The perinatal period is a high-risk time for onset of various psychiatric disorders. However, it is unclear how genetic risk factors for these disorders interact with biological changes associated with pregnancy and postpartum. This study evaluates whether psychiatric genome-wide association study (GWAS) results are enriched within various brain regions across the perinatal period.

**Methods:** Tissue-specific enrichment analyses were conducted to estimate the potential impact of GWAS loci on transcriptional changes in the brain across the perinatal period. GWAS summary statistics were obtained for 26 psychiatric phenotypes. RNA-sequencing data was acquired from four brain regions (hypothalamus, hippocampus, cerebellum, and neocortex) in mice at six timepoints (virgin, 14- and 16-days post-conception, and 1-, 3- and 10-days postpartum).

**Results:** Hippocampus and neocortex in the early postpartum period are significantly enriched (q-value < 0.05) for genetic variants associated with schizophrenia (SCZ), bipolar disorder (BD), depressive symptoms, and major depressive disorder with suicidal features. The most significant enrichment occurred in the neocortex for SCZ and BD, peaking at postpartum day 1 (SCZ p-value = 3.85 × 10^-8^; BD p-value = 6.65 × 10^-5^). In the hippocampus, BD and SCZ were enriched at postpartum day 1 (SCZ p-value = 3.27 × 10^-3^; BD p-value = 4.33 × 10^-3^). No enrichment was observed in cerebellum or hypothalamus for any of the psychiatric traits tested.

**Conclusions:** The results accord with previous epidemiological studies and provide context in which to interpret GWAS results. Understanding the burden of genetic variants across the perinatal period may help prioritise pathways underlying onset of psychiatric disorders outside of pregnancy and postpartum periods.

## Introduction

The perinatal period, which encompasses pregnancy and up to 12 months postpartum, represents a high-risk period for the onset of various psychiatric phenotypes. Yet underlying biological mechanisms remain inchoate, and research comprises largely epidemiological studies in hospital care settings, with low cohort diversity and little bio-psychosocial analyses (Kimmel et al., 2016; Meltzer-Brody et al., 2011; Munk-Olsen et al., 2006; Perry, Gordon-Smith, Di Florio, et al., 2021; Perry, Gordon-Smith, Jones, and Jones, 2021). Clinically, perinatal psychiatric symptoms can include depression, anxiety, and in rare cases, psychosis (L. M. Howard et al., 2014). Psychiatric disorders including eating disorders (EDs), bipolar disorder (BD), schizophrenia (SCZ), depression, and component traits have been observed to occur in the perinatal period pose specific threats to the well-being of the birth parent and offspring. The biological changes unique to the perinatal period may drive psychopathological mechanisms underpinned by genetic factors in a ‘gene × environment’ interaction. Onset often coincides with the extreme hormonal fluctuations characteristic of the perinatal period, that peak just before parturition and then fall precipitously. Additionally, morphological brain changes are associated with hormonal changes across the perinatal period (Chechko et al., 2022; Hoekzema et al., 2017; Sacher et al., 2020).

Outside of pregnancy, genetic and environmental factors contribute in a complex manner to psychiatric disorder aetiology. Recent genome-wide association studies (GWAS) have identified disorder-specific risk loci and show specific brain tissues and cell types are enriched for genetic variants contributing to heritability (Bryois et al., 2020; D. M. Howard et al., 2019; Trubetskoy et al., 2022). To our knowledge, psychiatric enrichment studies to date have been cross sectional in nature, broadly considering the adult human brain (Been et al., 2022; Paternina-Die et al., 2024; Servin-Barthet et al., 2025). The perinatal period is perhaps one of the most temporally influenced periods in the human life span, and an ideal context to examine how genomic variation can differentially affect a tissue or cell type over time. Genomic effects may be mediated by gene transcription, resulting in observed brain structural changes and even psychiatric symptoms.

Perinatal psychiatric symptom onset and structural brain changes may be mediated by gene transcription, in turn driven by temporal perinatal hormone fluctuations. Transcription patterns can in turn be altered by genetic variants (GTEx Consortium, 2020), increasing risk for various psychiatric disorders outside of the perinatal period. Enrichment with longitudinal perinatal brain expression data could preliminarily inform these questions and has not been performed, for either human or model organisms. Moreover, gene set pathway analyses for implicated genes presents one avenue into providing putative functional context for genetics risk expression and actionable hypotheses for functional investigation. Therefore, our goal was to perform a novel study to identify brain regions and perinatal time points showing enrichment for psychiatric GWAS results using perinatal mouse bulk-tissue RNA sequencing. We additionally aimed to perform gene set analysis for genes implicated in the enrichment analysis with freely available gene ontology databases. Linking genetic risk to psychiatric manifestation via nuanced Identifying genetically driven biological risk factors could in the long term inform personalised patient monitoring and therapeutics.

## Methods

European ancestry GWAS summary statistics were obtained for the following psychiatric phenotypes: anorexia nervosa (AN; Watson et al., 2019), BD (Mullins et al., 2021), BD subtypes I and II (BDI, BDII; Mullins et al., 2021), major depressive disorder (MDD; Wray et al., 2018), SCZ (Trubetskoy et al., 2022), depressive symptoms (D. M. Howard et al., 2019), treatment resistant disorders requiring electro-convulsive therapy (ECT; both general diagnoses and MDD-specific; Clements et al., 2021), MDD subtypes (Nguyen et al., 2022), and postpartum depression (PPD; Guintivano et al., 2023). All summary statistics were standardised to human genome build hg19 (using liftOver) with all RSIDs annotated to ENSEMBL GRCh37, Release 92 (Yates et al., 2016).

Raw bulk RNA-seq data used in this study were generated previously and described in full elsewhere (Ray et al., 2016). Briefly, four brain regions (hypothalamus, hippocampus, cerebellum, and neocortex) were acquired from C57BL/6J (B6) female mice. To capture perinatal temporal transcriptional changes, perinatal stages virgin, 14- and 16-days post conception (PC14 and PC16), and 1-, 3- and 10-days postpartum with nursing pups (PP1, PP3, and PP10) were used, with three biological replicates per timepoint (*N* total=72). For the purposes of this study, tissues were defined as ‘brain region × timepoint,’ for example, ‘hippocampus × PC14’ differs from ‘hippocampus × PP3’. Consequently, 24 different tissue types were examined. Calculations to convert the mouse timepoints to human equivalents were done as described in Agoston et al. (2017), where gestation in rats occurs approximately 12.7x faster, and weaning (postpartum) occurs approximately 8.6x faster comparative with humans. For example, the human equivalent of PC14 is ∼6.357 months/early third trimester, and the human equivalent of PP1 is ∼ 8.6 days/∼one 1 week.

To minimise potential error, an in-house bioinformatics pipeline was used to perform quality control, alignment, and expression quantification with the raw RNA data. Given the rapid development of bioinformatics tools, we created a pipeline with the most up-to-date versions of bioinformatics tools, catering to the type of RNA expression data being used and our aims, focusing on alignment sensitivity over speed (Sahraeian et al., 2017).

Raw RNA-seq data were obtained from NCBI Gene Expression Omnibus (accession GSE70732) and quality checked with FastQC (v0.11.9). Alignment was conducted with STAR (v2.7.10a; Dobin et al., 2012) to mouse reference genome GRCm38 (Ensembl build 107; human hg19 equivalent). Transcript quantification was performed for each sample using StringTie (v2.1.5; Pertea et al., 2015) to estimate transcripts per million (TPM) for each annotated gene.

TPM data was processed for enrichment analyses as reported previously (Bryois et al., 2020). Briefly, mean TPM was computed for each gene per tissue. Exclusion criteria included: non-unique names; no expression in any tissue; non-protein-coding genes; no 1:1 mouse-human orthologs; or occurrence in a region orthologous to the human MHC region (chr6, 25Mb to 34Mb). Orthologs between mice and humans were defined using the Compara database (Ensembl biomart). Gene expression was then scaled to a total of 1 million TPM per tissue. A gene expression specificity metric was calculated by dividing the expression of each gene in each tissue type by the total expression of that gene in all tissue types, leading to values ranging from 0-1 for each gene (0: no gene expression in that tissue; 1: 100% of that gene’s expression occurred in that tissue).

Using the 10% most specific genes per tissue, Jaccard clustering was used to identify overlap within tissues and across timepoints. Clustering indicated partial overlap within tissues and timepoints, and limited overlap across tissues, supporting the use of this threshold for enrichment analyses (Supplementary Figure 1). To further assess comparability of mouse data to human phenotypes, mean TPM of these tissue-specific genes were compared to mean TPM values for homologous genes in corresponding brain regions within Genotype-Tissue Expression data (GTEx); v8; GTEx Consortium, 2020). The association of mouse and human TPM data was assessed using linear regression models in R and were significantly associated (p < 2.2 × 10-16) in all four brain regions (Supplementary Figure 2).

Partitioned LD score regression (LDSC; Bulik-Sullivan et al., 2015) was used to test enrichment of tissue-specific genes for heritability in psychiatric traits. The genomic locations of the tissue-specific genes were mapped to human orthologs. Partitioned LDSC evaluates whether the per-SNP heritability differs for SNPs in the annotation (i.e., specific genes in a tissue) compared to SNPs that are not in the annotation, using GWAS summary statistics. Only genes with mean TPM ≥1 in each tissue were used for this analysis. SNPs located in 100kb windows (window size determined previously; Bryois et al., 2020) surrounding each of the tissue-specific genes were added to the baseline model (consisting of 53 different annotations provided by LDSC; Finucane et al., 2015) independently for each tissue. Next, coefficient Z-score p-values were used as a measure of tissue-trait association. Significance threshold was set to 5% false discovery rate (FDR) across all traits within each tissue.

Finally, gene set pathway analysis was performed for tissue-specific enriched genes identified in the enrichment analysis. Gene set analysis was performed in R using a hypergeometric test, with Gene Ontology terms (Ashburner et al., 2000; Gene Ontology Consortium et al., 2023). Results were visualised with rrvgo in R (Sayols, 2023). Output was filtered to contain the top five most FDR significant unique ‘gene set × tissue’ pairings in the most epidemiologically relevant tissues.

## Results

Across the major psychiatric diagnoses examined here (AN, BD, MDD, SCZ), significant enrichment (q-value < 0.05) was found for BD and SCZ in neocortex and hippocampus (Figure 1a). In neocortex, SCZ was enriched at all timepoints tested and BD was enriched for all but PC14. Additionally, patterns of enrichment were similar for BD and SCZ in neocortex, with enrichment significance increasing across pregnancy, peaking at PP1 (SCZ p-value = 3.85 × 10^-8^; BD p-value = 6.65 × 10^-5^), and declining for the remainder of the postpartum period. In the hippocampus, BD and SCZ were enriched at a single timepoint, PP1 (SCZ p-value = 3.27 × 10^-3^; BD p-value = 4.33 × 10^-3^). In the neocortex, AN trended towards the significance threshold at PC16 and PP1. No enrichment was observed in cerebellum or hypothalamus for any of the major psychiatric diagnoses tested. The full set of enrichment analysis results are in Supplementary Table 3.1.

**Figure 1:**
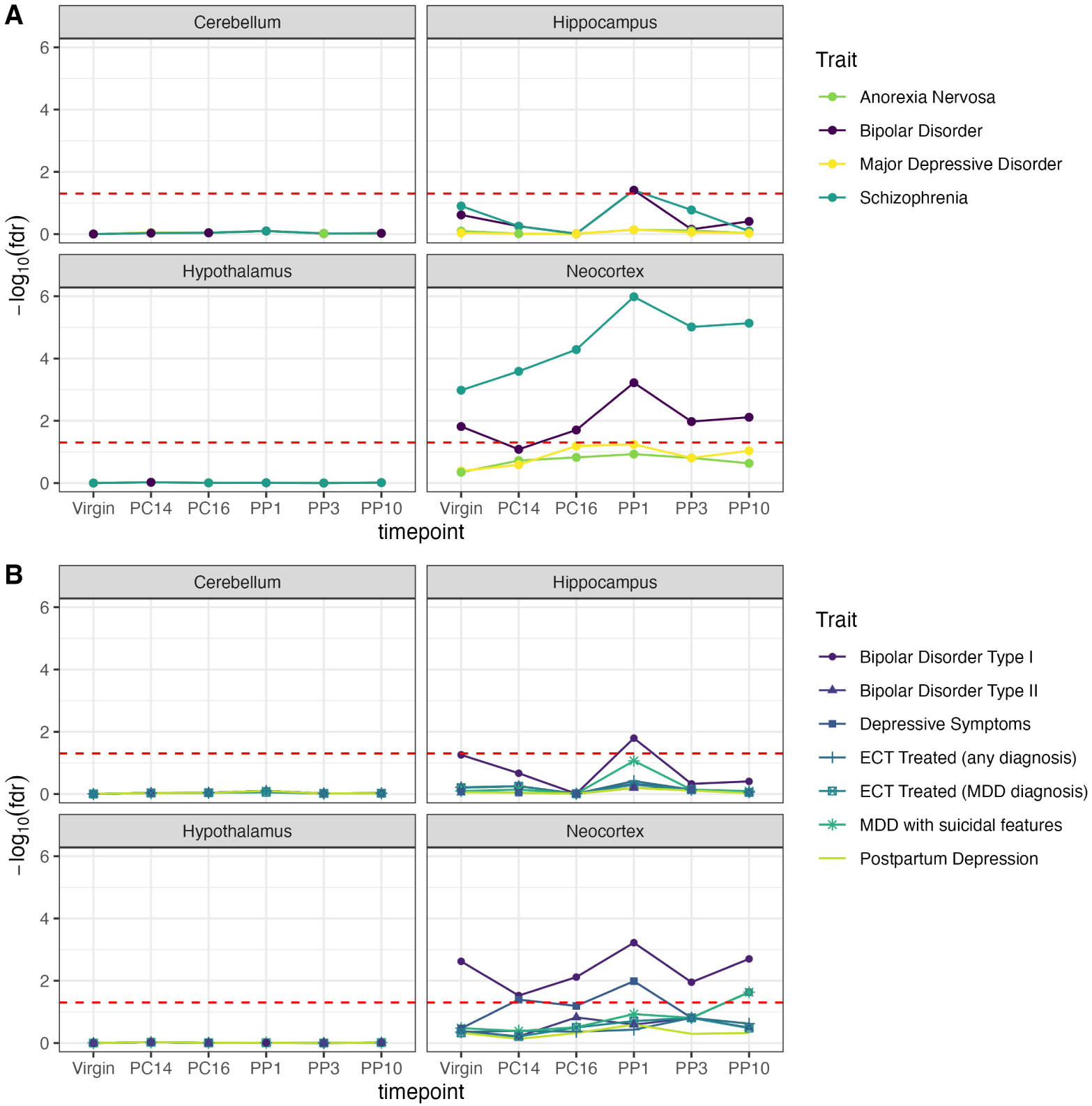
The y-axis is the -log10 of the false discovery rate (FDR), the x-axis is the six time points at which gene expression was measured, virgin, post conception day 14 (PC14; human equivalent = ∼6.357 months/early third trimester), post conception day 16 (PC16; human equivalent = ∼7.26 months/mid third trimester), postpartum day 1 (PP1; human equivalent = ∼ 8.6 days/∼one 1 week), postpartum day 3 (PP3; human equivalent = ∼25.8 days/∼3.5 weeks), and postpartum day (PP10; human equivalent = 86 days/∼12.3 weeks/∼three 3 months). The four panels are for each of the different brain regions, cerebellum (top left), hippocampus (top right), hypothalamus (bottom left), neocortex (bottom right). The red dotted line denotes the significance threshold. **A)** Each of the psychiatric disorders which were assessed with linkage disequilibrium score regression for enrichment in gene expression patterns is denoted by a different coloured line, the traits being anorexia nervosa, bipolar disorder, depression, and schizophrenia, listed in a key on the right-hand side of the figure. **B)** Each of the psychiatric trait phenotypes which were assessed with linkage disequilibrium score regression for enrichment in gene expression patterns is denoted by a different coloured line, the traits being bipolar disorder type I, bipolar disorder type II, electroconvulsive therapy for any diagnosis (ECT treated [any diagnosis]), electroconvulsive therapy for major depressive disorder diagnosis (ECT treated [MDD diagnosis]), major depressive disorder with suicidal features (MDD with suicidal features), and postpartum depression (Postpartum Depression), listed in a key on the right hand side of the figure.

Next, specific psychiatric subtypes and traits showed similar patterns of enrichment as BD and SCZ, and enrichment was limited to neocortex and hippocampus (Figure 1b). In neocortex, BDI was enriched at all timepoints, also peaking at PP1 (p-value = 6.65 × 10^-5^). Depressive symptoms showed significant enrichment in the neocortex at PP1 (p-value = 1.53 × 10^-3^), as well as at PP10 and PC14. Additionally, MDD with suicidal features showed enrichment in neocortex at PP10 (p-value = 4.33 × 10^-3^). In hippocampus, BDI was the only subtype that showed significant enrichment, at PP1 (p-value = 5.93 × 10^-4^).

Gene-set enrichment analysis for the significant tissues (neocortex virgin-PP10, and hippocampus PP1) resulted in 285 unique ‘gene set × tissue’ pairings (Supplementary Table 3.2). Table 1 shows the top 5 most FDR significant unique gene sets for each of the significant tissues (neocortex virgin through to PP10, and hippocampus PP1). The most significant gene sets implicated in three tissues of most epidemiological interest (hippocampus PP1, neocortex PP1, and neocortex PP3) were primarily involved in neuronal organisation and signalling, with several bio-behavioural responses in neocortex PP1.

**Table 1:** Unique gene sets for tissues with significant expression

| Annotation | Gene Set | Q-value | Overlap |
| --- | --- | --- | --- |
| HC PP1 | GO:0019725 GOBP: cellular homeostasis | 7.28x10<sup>-05</sup> | 93 |
| HC PP1 | GO:0050801 GOBP: ion homeostasis | 7.41x10<sup>-05</sup> | 84 |
| HC PP1 | GO:0032591 GOCC: dendritic spine membrane | 1.08x10<sup>-04</sup> | 8 |
| HC PP1 | GO:0006873 GOBP: cellular ion homeostasis | 1.50x10<sup>-04</sup> | 73 |
| HC PP1 | GO:0051402 GOBP: neuron apoptotic process | 1.54x10<sup>-04</sup> | 35 |
| NC Virgin | GO:0001216 GOMF: DNA binding transcription activator activity | 8.428x10<sup>-04</sup> | 53 |
| NC Virgin | GO:0017017 GOMF: map kinase tyrosine serine threonine phosphatase activity | 1.738x10<sup>-03</sup> | 6 |
| NC Virgin | GO:0045944 GOBP: positive regulation of transcription by RNA polymerase II | 3.625x10<sup>-03</sup> | 107 |
| NC Virgin | GO:0022030 GOBP: telencephalon glial cell migration | 4.819x10<sup>-03</sup> | 8 |
| NC Virgin | GO:0021892 GOBP: cerebral cortex GABAergic interneuron differentiation | 6.102x10<sup>-03</sup> | 5 |
| NC PC14 | GO:0071900 GOBP: regulation of protein serine threonine kinase activity | 9.501x10<sup>-03</sup> | 41 |
| NC PC14 | GO:0045859 GOBP: regulation of protein kinase activity | 9.621x10<sup>-03</sup> | 63 |
| NC PC14 | GO:0006182 GOBP: CGMP biosynthetic process | 1.127x10<sup>-02</sup> | 5 |
| NC PC14 | GO:0046068 GOBP: CGMP metabolic process | 1.128x10<sup>-02</sup> | 6 |
| NC PC14 | GO:0019207 GOMF: KINASE regulator activity | 1.571x10<sup>-02</sup> | 29 |
| NC PC16 | GO:0000149 GOMF: SNARE binding | 2.882x10<sup>-03</sup> | 19 |
| NC PC16 | GO:0060419 GOBP: heart growth | 4.679x10<sup>-03</sup> | 15 |
| NC PC16 | GO:0140238 GOBP: presynaptic endocytosis | 7.919x10<sup>-03</sup> | 12 |
| NC PC16 | GO:0097720 GOBP: calcineurin mediated signalling | 9.129x10<sup>-03</sup> | 11 |
| NC PC16 | GO:0140059 GOBP: dendrite arborization | 1.018x10<sup>-02</sup> | 5 |
| NC PP1 | GO:0001975 GOBP: response to amphetamine | 3.32x10<sup>-03</sup> | 9 |
| NC PP1 | GO:0001964 GOBP: startle response | 3.40x10<sup>-03</sup> | 8 |
| NC PP1 | GO:0042596 GOBP: fear response | 3.61x10<sup>-03</sup> | 10 |
| NC PP1 | GO:0150099 GOBP: neuron glial cell signalling | 4.37x10<sup>-03</sup> | 4 |
| NC PP1 | GO:0098926 GOBP: postsynaptic signal transduction | 5.42x10<sup>-03</sup> | 10 |
| NC PP3 | HP:0011890 HP: prolonged bleeding following procedure | 6.43x10<sup>-03</sup> | 7 |
| NC PP3 | GO:0062237 GOBP: protein localization to post synapse | 7.41x10<sup>-03</sup> | 9 |
| NC PP3 | HP:0004846 HP: prolonged bleeding after surgery | 7.41x10<sup>-03</sup> | 6 |
| NC PP3 | GO:0008289 GOMF: lipid binding | 8.19x10<sup>-03</sup> | 76 |
| NC PP3 | GO:0035255 GOMF: ionotropic glutamate receptor binding | 8.86x10<sup>-03</sup> | 7 |
| NC PP10 | GO:0010612 GOBP: regulation of cardiac muscle adaptation | 9.341x10<sup>-04</sup> | 6 |
| NC PP10 | GO:0021756 GOBP: striatum development | 2.211x10<sup>-03</sup> | 7 |
| NC PP10 | GO:0099626 voltage-gated calcium channel activity involved in regulation of presynaptic cytosolic calcium levels | 9.661x10<sup>-03</sup> | 4 |
| NC PP10 | GO:0014745 GOBP: negative regulation of muscle adaptation | 1.128x10<sup>-02</sup> | 5 |
| NC PP10 | HP:0001600 HP: abnormality of the larynx | 1.345x10<sup>-02</sup> | 24 |

The top five most FDR significant unique gene sets for each of the significant tissues, Hippocampus (HC) PP1, Neocortex (NC) virgin, Neocortex PC14, Neocortex PC16, Neocortex PP1, Neocortex PP3, and Neocortex PP10. Significance measure was false discovery rate (FDR) score for Gene Ontology (GO) parent term. GO identifiers and plain language term are given for the gene set (column 2). FDR threshold was set to q-value ;0.05. Overlap (column 4) is the number of genes overlapped between enrichment analysis and gene set analysis for that tissue.

## Discussion

Our data reflects epidemiologically observed risk patterns and highlights the temporality and tissue specificity of genetic risk for perinatal psychiatric disorders. The immediate postpartum period (approximately 10-19 days after childbirth) has the highest observed risk for psychiatric symptom onset within the perinatal period (Munk-Olsen et al., 2006). It is intriguing, therefore, that our findings show the most significant enrichment occurring one day postpartum in mice, approximately equivalent to one week postpartum in humans, and then at PP3, ∼3.5 weeks in humans (Agoston, 2017), indicating a potentially biological basis for this increased risk period (Figure 1). Identifying specific mechanisms of action underpinning this expression could potentially allow invaluable intervention methods at this critical period.

Schizophrenia, BD (specifically type 1), depressive symptoms, and MDD with suicidal features show significant enrichment across the perinatal period and reflect the most serious symptoms observed clinically. Incidentally, outside of the perinatal period, onset in women for all these disorders is most common in adolescence and early adulthood, which coincides with reproductive maturity and peak reproductive health. Women are more at risk of BD than men, and BD during the perinatal period carries with it high risk of both first-time psychosis and relapse (Conejo-Galindo et al., 2022; Di Florio et al., 2018). In an epidemiological review of BD postpartum relapse by Conejo-Galindo et al. (2022), the rate of BDI diagnosis comprised 69.83% of all BD relapses, although it was noted that differentiation of BD subtypes was heterogeneous across the total studies included for review. Genetic influences of SCZ, BD, and MDD with suicidal features may contribute to onset of postpartum psychosis and the high rates of perinatal suicide (Bergink et al., 2016; Conejo-Galindo et al., 2022; Jones et al., 2014). Conversely, the genetic influences of BD and depressive symptoms may contribute to the common occurrence of depression and anxiety observed in the perinatal period (Di Florio et al., 2013; Meltzer-Brody et al., 2018). Women who suffer BD prenatally face higher risk of postpartum relapse compared with other psychiatric disorders, including SCZ and MDD, and a perinatal episode of BD in the first pregnancy confers increased risk for recurrence as well as perinatal depression in subsequent pregnancies (Conejo-Galindo et al., 2022; Di Florio et al., 2018). Further, the increased significance of SCZ and BD compared to depressive symptoms may indicate a stronger genetic component for psychosis compared to depression more broadly. Identifying distinct, genetically driven biological risk mechanisms may be a key factor in helping differentiate pathological psychiatric indicators from the range of emotional responses inherent to such a significant life event.

Our analyses also provide insight into which brain regions in the perinatal period are most affected by genetic risk loci for multiple psychiatric traits and reflect broader neurobiological psychiatric findings (Ishida et al., 2023). Specifically, neocortex (primary centre for higher-order functioning; Paternina-Die et al., 2024), and hippocampus (primary centre for emotional regulation; Illouz et al., 2025; Sigurdsson and Duvarci, 2016) show significant enrichment for SCZ, BD, depressive symptoms, and MDD with suicidal features. Outside of a perinatal context, SCZ and BD loci show enrichment within these same regions (Mullins et al., 2021; Trubetskoy et al., 2022), while depressive symptoms only show enrichment in cortex (D. M. Howard et al., 2019). These findings are consistent with our results observed in virgin tissues, potentially reflecting the broader role of these brain regions in psychiatric illnesses. Moreover, the burden of genetic variants increases for psychiatric phenotypes (i.e. is higher in SCZ and BD compared with MDD) in the perinatal period, in the same pattern as prevalence for serious psychiatric illness more broadly (Munk-Olsen et al., 2006). The strong associations of neocortex and hippocampus further elucidate potential mechanisms for symptom onset at critical timepoints. Recent longitudinal evaluations of brain morphology across both pregnancy and postpartum, found that while cortical volume decreased during pregnancy, the direction of change reversed immediately postpartum, inversely mirroring the behaviour of hormones (Paternina-Die et al., 2024; Servin-Barthet et al., 2025). Additionally, changes could be discretised based on large scale functional networks, with the most grey matter loss and least recovery observed in the networks involved in higher order cognition, the default mode and frontoparietal networks (Servin-Barthet et al., 2025). This is especially when coupled with gene set analysis indicating enriched genes in these tissues are implicated in neuronal growth and differentiation processes (Table 1). Additionally, sex-specific hormones are known to have brain region specific effects in social behaviour (Been et al., 2022). Interestingly, gene set analysis for the genes associated with behaviour pathways are significantly expressed in the hippocampus at PP1. Cognitive and sensory disruptions in the cortex (Paternina-Die et al., 2024), along with mood disruptions from the hippocampus, may result in serious debilitating symptoms increasing risk for the parent and infant at a critical timepoint. Our data is an exciting progression towards delineating the biological mechanisms underpinning different perinatal psychiatric disorders and applying targeted and personalised therapeutics for the most efficacious outcomes.

Finally, the lack of enrichment for psychiatric traits tested (particularly AN and PPD) does not eliminate genetic contribution in onset of perinatal symptoms. Indeed, both disorders show notable heritability, and the metabo-psychiatric nature of AN in particular would be plausibly susceptible to the changes experienced in both these dimensions across the perinatal period. Heritability estimates indicate ∼50% of PPD phenotypic variability is attributable to genetics (Viktorin et al., 2016). As yet, genetic consisted primarily of candidate gene studies in small samples sizes that produce inconclusive results (Guintivano, Manuck, and Meltzer-Brody, 2018), and a single study done on genetic ancestry as a PPD predictor found no significant associations (Guintivano, Sullivan, et al., 2018). However, a recent large collaborative study found that perinatal depression had a higher polygenic risk score with MDD comparative with non-perinatal depression, indicating both a high severity and shared underlying genetic architecture (Kiewa et al., 2022). Perinatal depression also had significant PGS with SCZ, BD, AN, attention deficit hyperactivity disorder, and neuroticism (Kiewa et al., 2022). As global collaborations grow, so too do cohorts’ size and diversity, and consequently power to detect nuanced genetic architecture underpinning the disorder.Moreover, women in whom ED symptomology worsens during the postpartum period have a reported increased risk of PPD (Janas-Kozik et al., 2021; Meltzer-Brody et al., 2011). This lack may be due to differences in statistical power across summary statistics. Enrichment analyses can be performed again as updated analyses are released.

## Limitations and Future Considerations

This work builds on existing methods and datasets to link GWAS results to more precise biologically driven therapeutic practices (Bryois et al., 2020; Bulik-Sullivan et al., 2015; Finucane et al., 2015). However, there are some caveats. First, the mouse RNA-seq data comes from heterogeneous bulk tissues, prohibiting observation of cell-type specific changes. Additionally, the specificity metric used is influenced by the number of tissues assayed. As in other studies using this approach, the results should be interpreted cautiously. However, the use of mouse data is comparable to what is observed in human data (Supplementary Figure 3.2) and allows for interrogation of complex biology in brain tissues across multiple perinatal timepoints otherwise unfeasible in humans.

Another consideration is that the available GWAS results include male and female samples at differing ratios. Consequently, the observed patterns of enrichment are potentially diluted by the presence of males in the analysis of the perinatal period. This seems unlikely given the high genetic correlations between sexes (Martin et al., 2018; Nievergelt et al., 2019). Additionally, the summary statistics comprise only participants of European ancestry. Further work is needed to confirm the findings in populations of non-European ancestry.

Differentiating between brain regions improves granularity over bulk tissue gene expression, however, research increasingly shows the minutiae of brain function, with gene expression and regulatory processes differing even to the single nuclei level (Siletti et al., 2023). Studies using single cell or single nuclei brain expression data have shown notable improvement in extrapolating functional activities of psychiatric GWAS loci, and should be considered for functional validation of this study (Yao et al., 2025).

## Conclusions

These results highlight the perinatal period, particularly the immediate postpartum period, as a vulnerable window for genetic risk for several severe phenotypes including SCZ, BD (specifically type 1), depressive symptoms, and MDD with suicidal features. The findings converge on the same timepoints as previous epidemiological studies and underscore the context-specific nature for interpreting GWAS results of psychiatric disorders. We highlight again the novelty and importance our study, examining temporal perinatal brain expression. Biologically informed temporal genetic risk targets present a powerful and necessary avenue for perinatal psychiatric therapeutics.

## Data Availability

All data used in this study are free and publicly available. Raw RNA-seq data were obtained from NCBI Gene Expression Omnibus (accession GSE70732). European ancestry GWAS summary statistics were obtained from the PGC website (https://pgc.unc.edu/for-researchers/download-results/). Gene ontology terms were downloaded from the Gene Ontology database (geneontology.org).

## Acknowledgments

J Guintivano would like to acknowledge National Institute of Mental Health grant K01MH116413. N Berthold would like to acknowledge a fellowship awarded by the Centre of Excellence for Eating Disorders University of North Carolina at Chapel Hill, the Quad Fellowship awarded by the Institute of International Education, and an Australian Research Training Scholarship provided by the University of Western Australia.

## Author Contributions

NB contributed to concept development, study design, data analysis and manuscript preparation and editing. JG contributed to concept development, study design, data analysis and manuscript preparation and editing. SY contributed to data analysis, manuscript preparation and editing.

## Supplementary Materials

**Figure 2:**
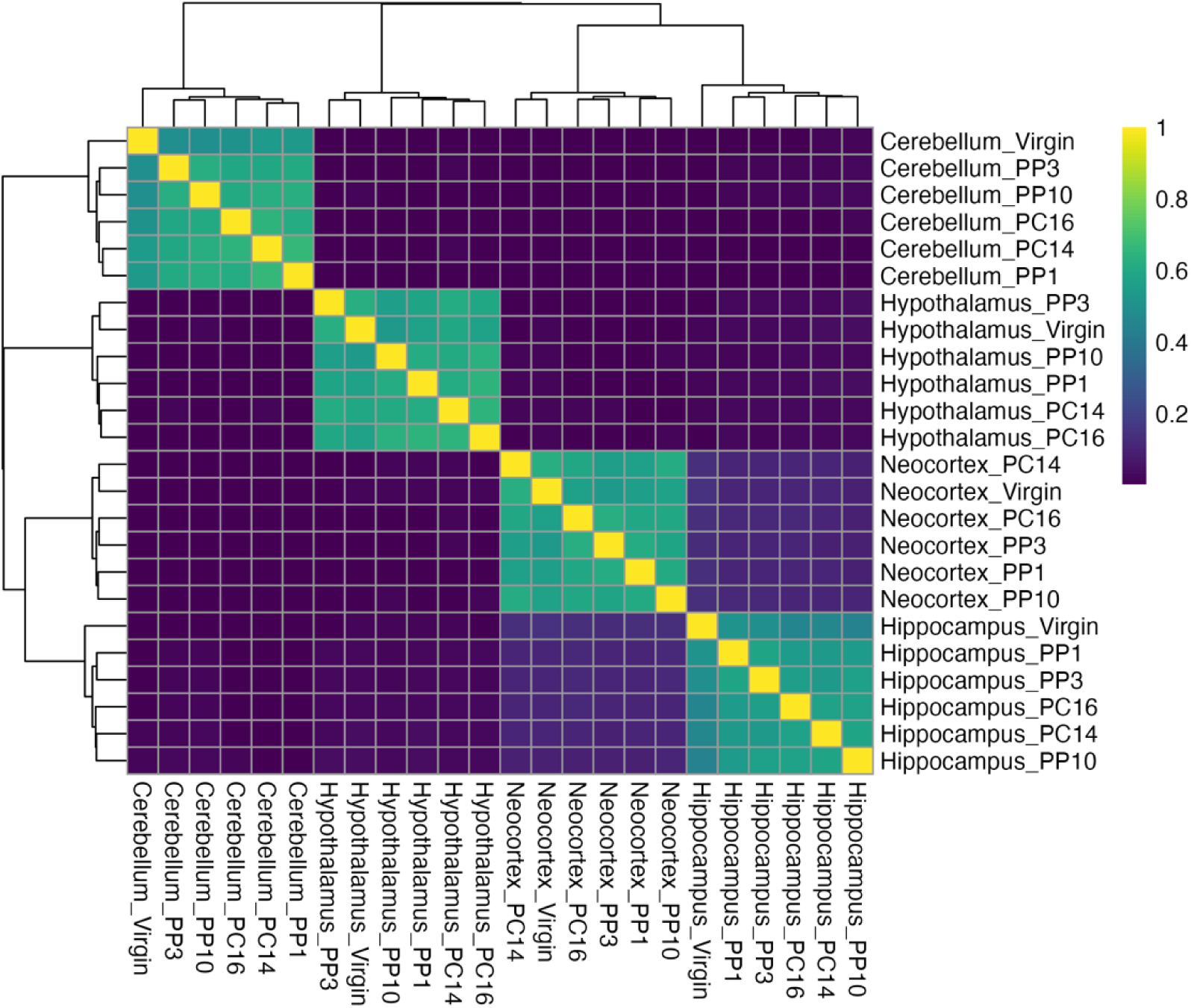
Jaccard index calculated for the similarity of tissue expression across time and between tissues. Scale between 0-1, where 0 is no similarity, and 1 is perfect similarity. Expression similarity occurs within brain regions, but not across time.

**Figure 3:**
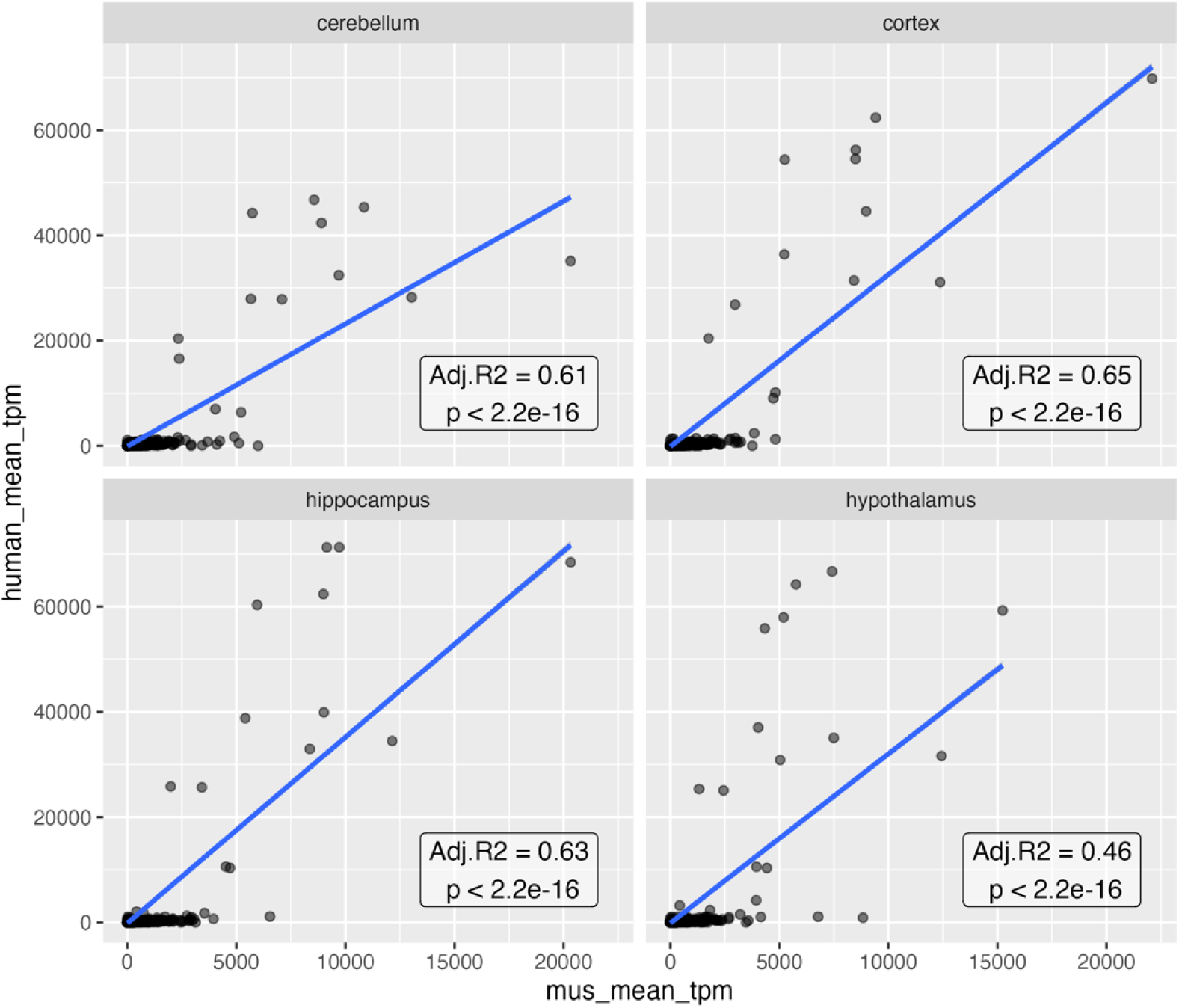
Correlation graphs produced by R showing the correlation of expression in the four brain regions (upper left = cerebellum, upper right = cortex, lower left = hippocampus, lower right = hypothalamus) between human tissues acquired from GTEx (y axis) and mouse tissues (x axis). Expression was significantly correlated (p¡0.05).

